# Chest wall abnormalities in Swiss childhood cancer survivors

**DOI:** 10.1101/2021.01.24.21250393

**Authors:** Rahel Kasteler, Christa Lichtensteiger, Christina Schindera, Marc Ansari, Claudia E. Kuehni, for the Swiss Pediatric Oncology Group (SPOG)

## Abstract

**Background:** Chest wall abnormalities are a poorly studied complication after treatment for childhood cancer. Chest wall abnormalities are not well described in the literature, and little is known on the impact on daily life of survivors.

**Methods:** We investigated chest wall abnormalities in the nationwide, population-based cohort study (Swiss Childhood Cancer Survivor Study) with a questionnaire survey to describe prevalence and risk factors. We then interviewed a nested sample of survivors to understand types of chest wall abnormalities and their impact on daily life of survivors.

**Results:** 48 of 2,382 (95%CI 2%–3%) survivors reported a chest wall abnormality. Risk factors were older age at cancer diagnosis (16–20 years; OR 2.5, 95%CI 1.0– 6.1), lymphoma (OR 3.8, 95%CI 1.2–11.4), and central nervous system tumors (OR 9.5, 95%CI 3.0–30.1) as underlying disease, and treatment with thoracic radiotherapy (OR 2.0, 95%CI 1.0–4.2), surgery to the chest (OR 4.5, 95%CI 1.8–11.5), or chemotherapy (OR 2.9, 95%CI 1.0–8.1) .The nature of the chest wall abnormalities varied and included thoracic wall deformities (30%), deformations of the spine (5%) or both (55%), and scars (10%). Chest wall abnormalities affected the daily life in two thirds (13/20) of those who reported these problems, and 15 (75%) had required chest wall abnormalities-related medical attention.

**Conclusion:** It is important that during follow-up care physicians pay attention to chest wall abnormalities, which are rare late-effect of cancer treatment, but can considerably affect well-being of cancer survivors.

## 1. Background

Chest wall abnormalities have been reported in a widely variable proportion of childhood cancer survivors, but available studies were small, including 16 to 143 participants,^1-9^ or focused on patients with selected cancer diagnoses like chest wall sarcoma,^1^ central nervous system (CNS) tumours,^10^ neuroblastoma^2,9^ or Wilms tumour.^3,4^ Some studies reported on specific cancer treatments like thoracic radiotherapy, radiotherapy to the spine,^6^ abdominal radiotherapy,^7,8^ or surgical interventions for solid tumours.^5^ Such data are not representative for the entire population of childhood cancer survivors. In large, cross-sectional studies, 1.4%^11^ (North America, multicenter study) to 2.0%^12^ (Switzerland, population based study) of survivors reported chest wall abnormalities in questionnaire surveys.^10-12^ Those investigations were questionnaire-based and used one single question on chest wall abnormalities. This did not allow to understand the nature of chest wall abnormalities as no exact definition was attached to the question. Chest wall abnormalities could include thoracic wall and spinal deformities, breast asymmetries, or scars, and the term may not have been clear to some participants. No study investigated whether and how chest wall abnormalities affect the daily life of survivors and if medical care is needed.

We approached these uncertainties by assessing the prevalence of chest wall abnormalities reported by survivors in Switzerland and investigating risk factors. We also interviewed a nested sample of survivors to understand the nature of the reported chest wall abnormalities and impact on daily life. Finally, we conducted a systematic review of the available literature on chest wall abnormalities in childhood cancer survivors.

## 2. Methods

### Swiss Childhood Cancer Survivor Study

The Swiss Childhood Cancer Survivor Study (SCCSS) is a population-based, long-term follow-up study of patients registered in the Swiss Childhood Cancer Registry (SCCR, www.childhoodcancerregistry.ch). Participants had been diagnosed with leukemia, lymphoma, CNS tumors, malignant solid tumors, or Langerhans cell histiocytosis since 1976 and before the age of 21 years. Participants, who had survived ≥5 years since initial cancer diagnosis and were alive at the time of the study,^13,14^ received a questionnaire between 2007 and 2013. Nonresponders received a second copy of the questionnaire four to six weeks later. If they again did not answer, we contacted them by phone. In total 2,382 survivors replied (Supplementary figure S1). Detailed methods of the SCCSS have been published.^12-14^

### Outcome: Chest wall abnormalities

The SCCSS questionnaire, like the North Amercian^15^ and British^16^ Childhood Cancer Survivor Studies, includes one question on chest wall abnormalities in the section on pulmonary health: *“Have you ever been told by a doctor that you have or have had changes on your thorax and/or ribs?”* and possible answers included *ever in live* (yes/no), *since when (year), currently* (yes/no) (Supplementary figure S2).

### Validation of outcome by telephone interview

In a nested follow-up study, we sent a letter to all survivors who had reported a chest wall abnormality in the questionnaire to invite them to take part in a telephone interview. All those were at least 18 years old, still alive, had consented to participation in further studies and had a valid telephone number. Survivors were contacted by telephone between July 2017 and September 2017 by one investigator (CL) (Supplementary figure S1). The purpose of the structured interview was to determine the medical problems underlying the chest wall abnormalities. We sought information on 1) deformations of the chest wall that included asymmetric chest wall, pectus excavatum, pectus carinatum, completely or partially missing ribs, deformation of the breast, muscular abnormalities or other deformations of the chest wall or ribs, 2) deformations of the spine including scoliosis, hyperkyphosis, hyperlordosis or other deformations of the spine, and 3) scars on the chest wall. We also asked about the impact of chest wall abnormalities on daily life which could include general complaints as well as cosmetic problems, respiration problems, flexibility impairments, pain because of the chest wall abnormalities, and impairment in activities of daily living. We also asked whether medical attention – consultation with a physician, diagnostic investigations, operations and physiotherapy – had been sought. The questionnaire used for the interview is available on request.

The Ethics Committee of the Canton of Bern approved the SCCR and the SCCSS (KEK-BE: 166/2014), and the Swiss Childhood Cancer Survivor Study is registered at ClinicalTrials.gov (identifier: NCT03297034).

### Covariates: Demographic and cancer-related characteristics

We obtained cancer characteristics from the SCCR including age at diagnosis, year of diagnosis, cancer diagnosis according to the International Classification of Childhood Cancer, 3rd edition ^17^, and details on radiotherapy, surgery, and chemotherapy. We combined total body irradiation, mantle field radiation, and radiation to the thorax, lungs, mediastinum, or thoracic spine to thoracic radiotherapy (yes/no). Surgery to the chest (yes/no) included the clavicles, scapulae and ribs, tumor excision from soft tissue on thorax, thoracic muscles, thoracic spine, and tumor or lymph node biopsies on the chest wall.

### Statistical analysis

We reported prevalence of chest wall abnormalities overall and stratified by sex, age at cancer diagnosis (0–5, 6–10, 11–15, 16–20), years of diagnosis (1976–1990, 1991–2005), cancer diagnosis (leukemia, lymphoma, CNS tumor, other tumors) and cancer treatment (thoracic radiotherapy (yes, no), surgery to the chest (yes, no), any chemotherapy (yes, no)). We identified demographic and cancer-related risk factors for chest wall abnormalities using univariable and multivariable logistic regression. All analyses were done in Stata (Version 14; Stata Corporation, Austin, TX).

### Systematic literature review

We conducted a literature review searching for relevant articles in the two bibliographic databases PubMed and Embase Ovid, last updated December 31th 2019. Both databases were searched using thesaurus terms (MeSH, Emtree) and textwords. We applied restrictions to language and searched studies on humans only, and excluded from the search conference abstracts, letters to the editor and editorials. To retrieve further relevant publications, we checked the reference lists of studies included and added Google scholar for a full-text search. An information specialist from the University Library of Bern was consulted to set up the search strategies in order to ensure optimal data acquisition.

The search results were screened in two steps by two independent reviewers (RK, CL) and assessed according to relevance and eligibility criteria (PRISMA flow diagram). Additional articles were searched by screening the reference list of suitable systematic reviews found in the two databases. For details on search strategies and search platforms, see Supplementary Text and Supplementary figure S3.

## 3. Results

### Prevalence and risk factors of chest wall abnormalities

Among 2,382 survivors who participated in the SCCSS, 2% (48/2,382) reported a chest wall abnormality (95% confidence interval [95%CI] 1.5–2.7). Male survivors were more often affected (2.5%, 95%CI 1.8–3.5) than females (1.4%, 95%CI 0.8–2.3) (table 1). Median age at study was 31 years (interquartile range [IQR] 25 – 38) and survivors who were older at cancer diagnosis (16-20 years, 4.2%, 95%CI 2.2 – 7.9) had a higher prevalence of chest wall abnormalities compared to those who were younger. Prevalence did not change over time, being 2% and 2.1% in the periods of 1976–1990 and 1991–2005. When comparing underlying diagnoses, chest wall abnormalities were most frequently reported by survivors of lymphoma (3.7%, 95%CI 2.3–6.0) and CNS tumors (3.8%, 95%CI 2.2–6.4), but rarely by participants treated for leukemia (0.6%, 95%CI 0.2–1.5). Nearly 8% of survivors treated with surgery to the chest and 6% treated with thoracic radiotherapy reported a chest wall abnormality.

**TABLE 1.**
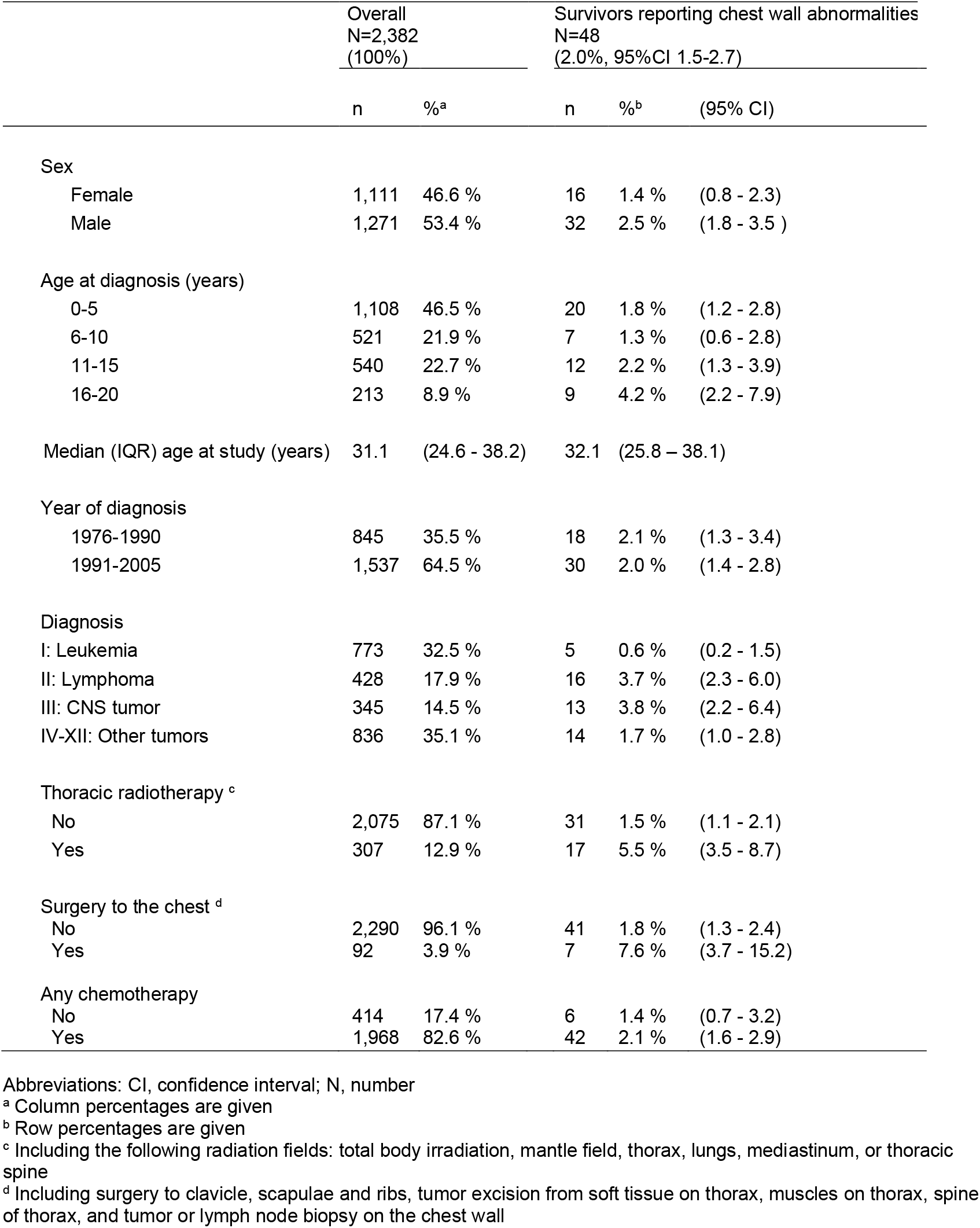
Characteristics of Swiss childhood cancer survivors overall and of those reporting chest wall abnormalities.

|  | Overall<br>N=2,382<br>(100%) |  | Survivors reporting chest wall abnormalities<br>N=48<br>(2.0%, 95%CI 1.5-2.7) |  |  |
| --- | --- | --- | --- | --- | --- |
|  | n | %<sup>a</sup> | n | %<sup>b</sup> | (95% CI) |
| Sex |  |  |  |  |  |
| Female | 1,111 | 46.6 % | 16 | 1.4 % | (0.8 - 2.3) |
| Male | 1,271 | 53.4 % | 32 | 2.5 % | (1.8 - 3.5 ) |
| Age at diagnosis (years) |  |  |  |  |  |
| 0-5 | 1,108 | 46.5 % | 20 | 1.8 % | (1.2 - 2.8) |
| 6-10 | 521 | 21.9 % | 7 | 1.3 % | (0.6 - 2.8) |
| 11-15 | 540 | 22.7 % | 12 | 2.2 % | (1.3 - 3.9) |
| 16-20 | 213 | 8.9 % | 9 | 4.2 % | (2.2 - 7.9) |
| Median (IQR) age at study (years) | 31.1 | (24.6 - 38.2) | 32.1 | (25.8 – 38.1) |  |
| Year of diagnosis |  |  |  |  |  |
| 1976-1990 | 845 | 35.5 % | 18 | 2.1 % | (1.3 - 3.4) |
| 1991-2005 | 1,537 | 64.5 % | 30 | 2.0 % | (1.4 - 2.8) |
| Diagnosis |  |  |  |  |  |
| I: Leukemia | 773 | 32.5 % | 5 | 0.6 % | (0.2 - 1.5) |
| II: Lymphoma | 428 | 17.9 % | 16 | 3.7 % | (2.3 - 6.0) |
| III: CNS tumor | 345 | 14.5 % | 13 | 3.8 % | (2.2 - 6.4) |
| IV-XII: Other tumors | 836 | 35.1 % | 14 | 1.7 % | (1.0 - 2.8) |
| Thoracic radiotherapy <sup>c</sup> |  |  |  |  |  |
| No | 2,075 | 87.1 % | 31 | 1.5 % | (1.1 - 2.1) |
| Yes | 307 | 12.9 % | 17 | 5.5 % | (3.5 - 8.7) |
| Surgery to the chest <sup>d</sup> |  |  |  |  |  |
| No | 2,290 | 96.1 % | 41 | 1.8 % | (1.3 - 2.4) |
| Yes | 92 | 3.9 % | 7 | 7.6 % | (3.7 - 15.2) |
| Any chemotherapy |  |  |  |  |  |
| No | 414 | 17.4 % | 6 | 1.4 % | (0.7 - 3.2) |
| Yes | 1,968 | 82.6 % | 42 | 2.1 % | (1.6 - 2.9) |

Abbreviations: CI, confidence interval; N, number
<sup>a</sup> Column percentages are given
<sup>b</sup> Row percentages are given
<sup>c</sup> Including the following radiation fields: total body irradiation, mantle field, thorax, lungs, mediastinum, or thoracic spine
<sup>d</sup> Including surgery to clavicle, scapulae and ribs, tumor excision from soft tissue on thorax, muscles on thorax, spine of thorax, and tumor or lymph node biopsy on the chest wall

In a multivariable regression, the following factors remained independently associated with chest wall abnormalities: male sex (odds ratio [OR] 1.8, 95%CI 1.0–3.3), older age at cancer diagnosis (OR 2.5, 95%CI 1.0–6.1), lymphoma (OR 3.8, 95%CI 1.2–11.4), CNS tumor (OR 9.5, 95%CI 3.0–30.1), thoracic radiotherapy (OR 2.0, 95%CI 1.0–4.2), surgery to the chest (OR 4.5, 95%CI 1.8–11.5), and chemotherapy (OR 2.9, 95%CI 1.0–8.1) (table 2).

**TABLE 2.** Demographic and cancer-related risk factors for chest wall abnormalities in Swiss childhood cancer survivors.

| Total N=2,382 | Chest wall abnormalities (n=48) |  |  |  |  |  |
| --- | --- | --- | --- | --- | --- | --- |
|  | OR<sub>crude</sub><sup>a</sup> | (95%CI) | P<sup>b</sup> | OR<sub>adj</sub><sup>c</sup> | (95%CI) | P<sup>c</sup> |
| Sex |  |  | **0.059** |  |  | **0.062** |
| Female | Ref. |  |  | Ref. |  |  |
| Male | 1.8 | (1.0–3.2) |  | 1.8 | (1.0–3.3) |  |
| Age at diagnosis (years) |  |  | **0.125** |  |  | **0.017** |
| 0-5 | Ref. |  |  | Ref. |  |  |
| 6-10 | 0.7 | (0.3–1.8) |  | 0.5 | (0.2–1.2) |  |
| 11-15 | 1.2 | (0.6–2.5) |  | 0.7 | (0.3–1.6) |  |
| 16-20 | 2.4 | (1.1–5.3) |  | 2.5 | (1.0–6.1) |  |
| Year of diagnosis |  |  | **0.768** |  |  | **0.602** |
| 1976-1990 | Ref. |  |  | Ref. |  |  |
| 1991-2005 | 0.9 | (0.5–1.7) |  | 0.8 | (0.5–1.6) |  |
| Diagnosis |  |  | **<0.001** |  |  | **<0.001** |
| I: Leukemia | Ref. |  |  | Ref. |  |  |
| II: Lymphoma | 6.0 | (2.2–16.4) |  | 3.8 | (1.2–11.4) |  |
| III: CNS tumor | 6.0 | (2.1–17.0) |  | 9.5 | (3.0–30.1) |  |
| IV-XII: Other tumors | 2.6 | (0.9–7.3) |  | 2.1 | (0.7–6.1) |  |
| Thoracic radiotherapy <sup>d</sup> |  |  | **<0.001** |  |  | **0.058** |
| No | Ref. |  |  | Ref. |  |  |
| Yes | 3.9 | (2.1–7.1) |  | 2.0 | (1.0–4.2) |  |
| Surgery to the chest <sup>e</sup> |  |  | **0.002** |  |  | **0.004** |
| No | Ref. |  |  | Ref. |  |  |
| Yes | 4.5 | (2.0–10.4) |  | 4.5 | (1.8–11.5) |  |
| Any chemotherapy |  |  | **0.348** |  |  | **0.029** |
| No | Ref. |  |  | Ref. |  |  |
| Yes | 1.5 | (0.6–3.5) |  | 2.9 | (1.0–8.1) |  |

Abbreviations: CI, confidence interval; OR, Odds ratio; N, number; P, P-value
<sup>a</sup> Odds ratio from univariable logistic regression analysis
<sup>b</sup> P-values calculated from likelihood-ratio tests comparing survivors with and without chest wall abnormality
<sup>c</sup> Odds ratio from multivariable logistic regression analysis, model adjusted for all factors shown
<sup>d</sup> Including the following radiation fields: total body irradiation, mantle field, thorax, lungs, mediastinum, or thoracic spine
<sup>e</sup> Including surgery to clavicle, scapulae and ribs, tumor excision from soft tissue on thorax, muscles on thorax, spine of thorax, and tumor or lymph node biopsy on the chest wall

### Telephone interviews

Among the 48 survivors who reported chest wall abnormalities, 25 survivors were eligible for being interviewed and 20 participated (80%), 18 confirmed to have a chest wall abnormality (Table 3). When asked in more detail, 85% (17/20) described thoracic wall deformities, 60% (12/20) a deformation of the spine, and 70% (14/20) scars on the chest wall. Most survivors (80%;16/20) reported multiple problems (Figure 1 and Table 3). Thoracic wall deformities included pectus excavatum, pectus carinatum and unspecified thoracic asymmetries, missing or deformed ribs, and deformation of the breasts. Deformation of the spine included scoliosis, hyper kyphosis, and hyper lordosis.

**TABLE 3.** Impact of chest wall abnormalities on the daily life of survivors, and medical attention required because of chest wall abnormality.

|  | **N = 20** | **Proportion (%)** |
| --- | --- | --- |
| **Impact of chest wall abnormalities on the daily life** |  |  |
| Any impact | 13 | 65% |
| Respiration | 8 | 40% |
| Flexibility | 7 | 35% |
| Activities of daily living | 6 | 30% |
| Cosmetic | 6 | 30% |
| Pain | 6 | 30% |
| **Required medical attention because of chest wall abnormality** |  |  |
| Any medical attention | 15 | 75% |
| Consultation <sup>a</sup> | 14 | 70% |
| 1 specialist visited | 10 | 50% |
| >1 specialist visited | 4 | 20% |
| Diagnostic investigation <sup>b</sup> | 9 | 45% |
| Chest X-ray | 6 | 30% |
| Chest X-ray + lung function test | 3 | 15% |
| Operations | 1 | 5% |
| Physiotherapy | 6 | 30 |

Abbreviations: N, number; Respiration, any respiratory impairment; Flexibility, impairment of flexibility; Activities of daily living, inability or problems when performing activities of daily living such as housekeeping; Cosmetic, disturbed by the cosmetic appearance of the chest wall abnormality; Pain, any pain because of the chest wall abnormality; Consultation, ever consulted a medical doctor because of the chest wall abnormality; Diagnostic investigation, had further diagnostic testing because of the chest wall abnormality (e.g., chest x-ray, lung function tests); Operations, had an operation because of the chest wall abnormality; Physiotherapy, visited physical therapy because of the chest wall abnormalities
<sup>a</sup> Survivors reported consultations with: general practitioners n=5, pediatric oncologists n=4, oncologist n=1, orthopedist n=3, chiropractic's n=1, surgeons n=1, sports physicians n=1, rheumatologist n=1

**Figure 1.**
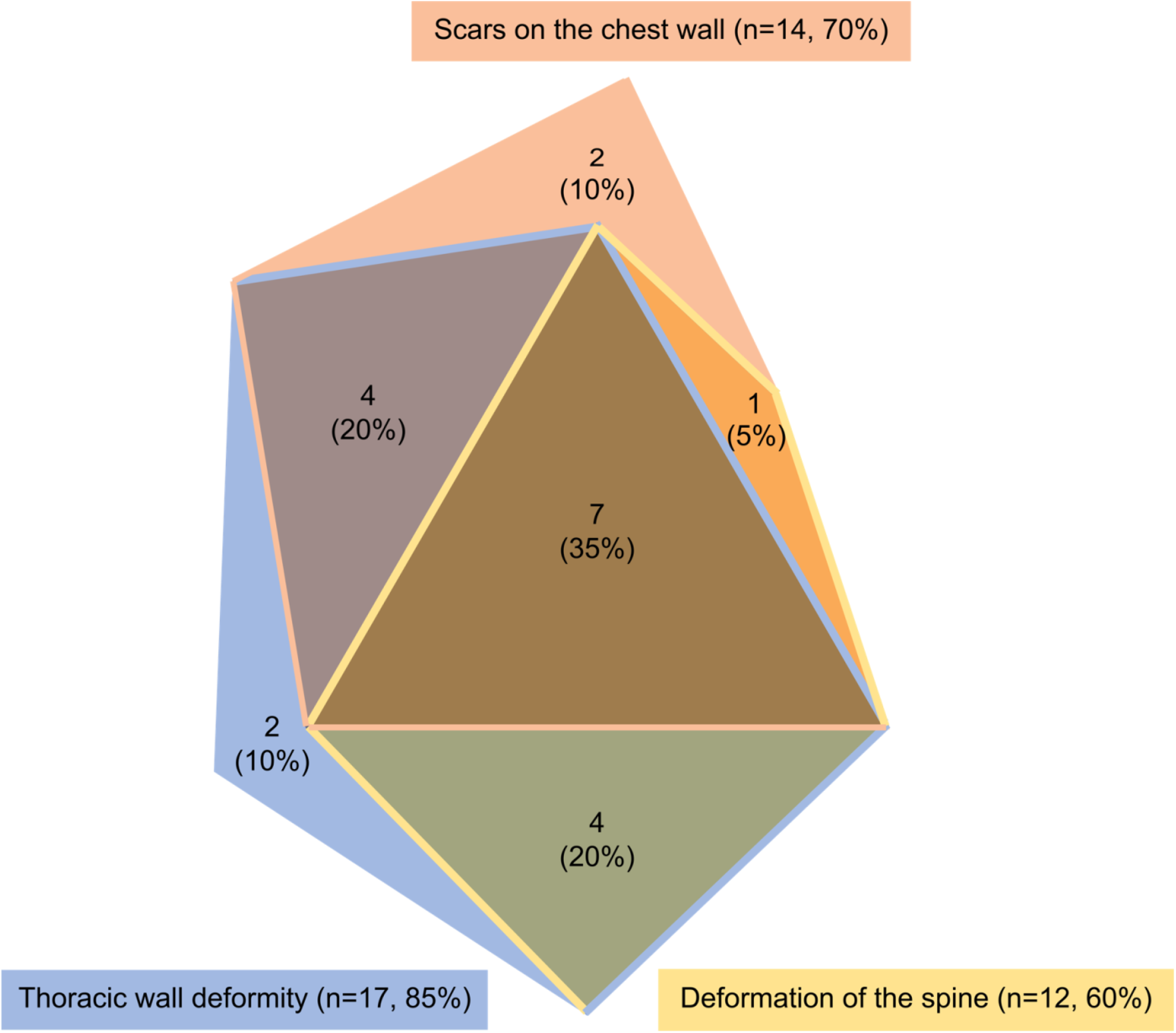
Proportional Venn-diagram showing reported types and overlap of chest wall abnormalities in childhood cancer survivors in the telephone interview.

We also asked survivors whether the chest wall abnormalities affected their daily life and if they had sought medical attention. Among the 20 survivors interviewed, 13 survivors said that the chest wall abnormality affected respiration, flexibility, and activities of daily life, caused cosmetic problems, or pain (table 3). Fifteen survivors had sought medical attention. Fourteen consulted a doctor (10 visited one specialist only and four visited multiple specialists). Specialists included general practitioners, pediatric oncologists, orthopedists, chiropractors, surgeons, sport physicians or rheumatologists. Nine survivors underwent diagnostic testing, six had a chest x-ray only and three a chest x-rays and lung function tests. One survivor had needed an operation, and six had been treated by physiotherapists (table 3).

### Literature Review

Of 2,167 potentially relevant articles identified, we excluded 1,935 articles after screening of title and abstracts, leaving 244 articles for full text screening. Of those, we excluded a further 232 articles that did not meet inclusion criteria (Supplementary figure S3). We summarized the remaining in table 4. Among these, only two investigated in an unselected cohort of survivors with regard to cancer diagnosis and treatment.^11,12^ Both used postal questionnaires (the Swiss and North American Childhood Cancer Survivor Studies) and found a prevalence of chest wall abnormalities of 2.0%^12^ and 1.3%^11^. Six studies focused on survivors of selected cancer diagnoses; one on chest wall sarcoma,^1^ one on CNS tumours,^10^ two on neuroblastoma,^2,9^ and two on Wilms tumour.^3,4^ Four studies focused on specific cancer treatments only: one on operations of solid tumours^5^ and three on irradiation including at least parts of the thorax^6-8^.

**Table 4.**
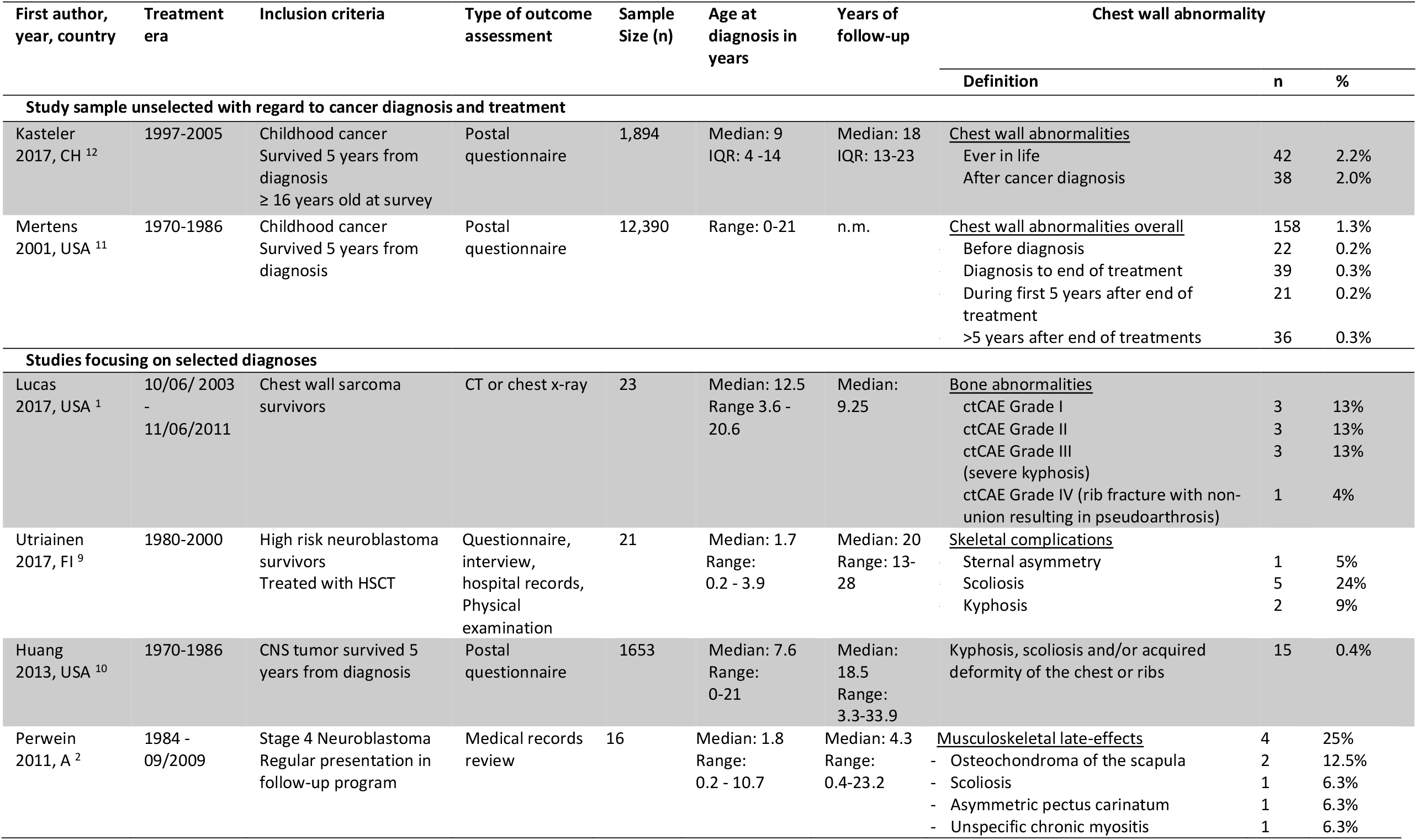
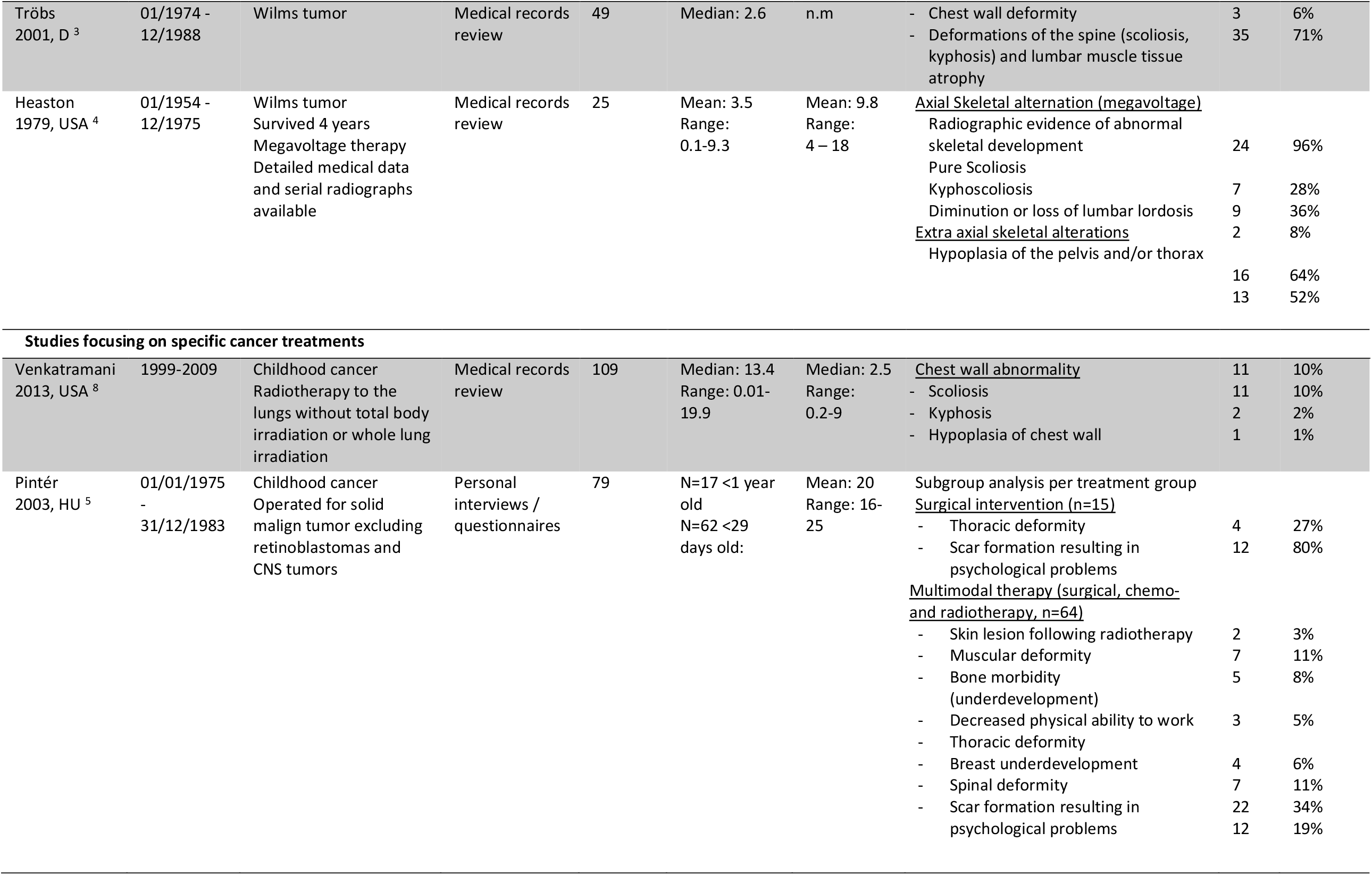
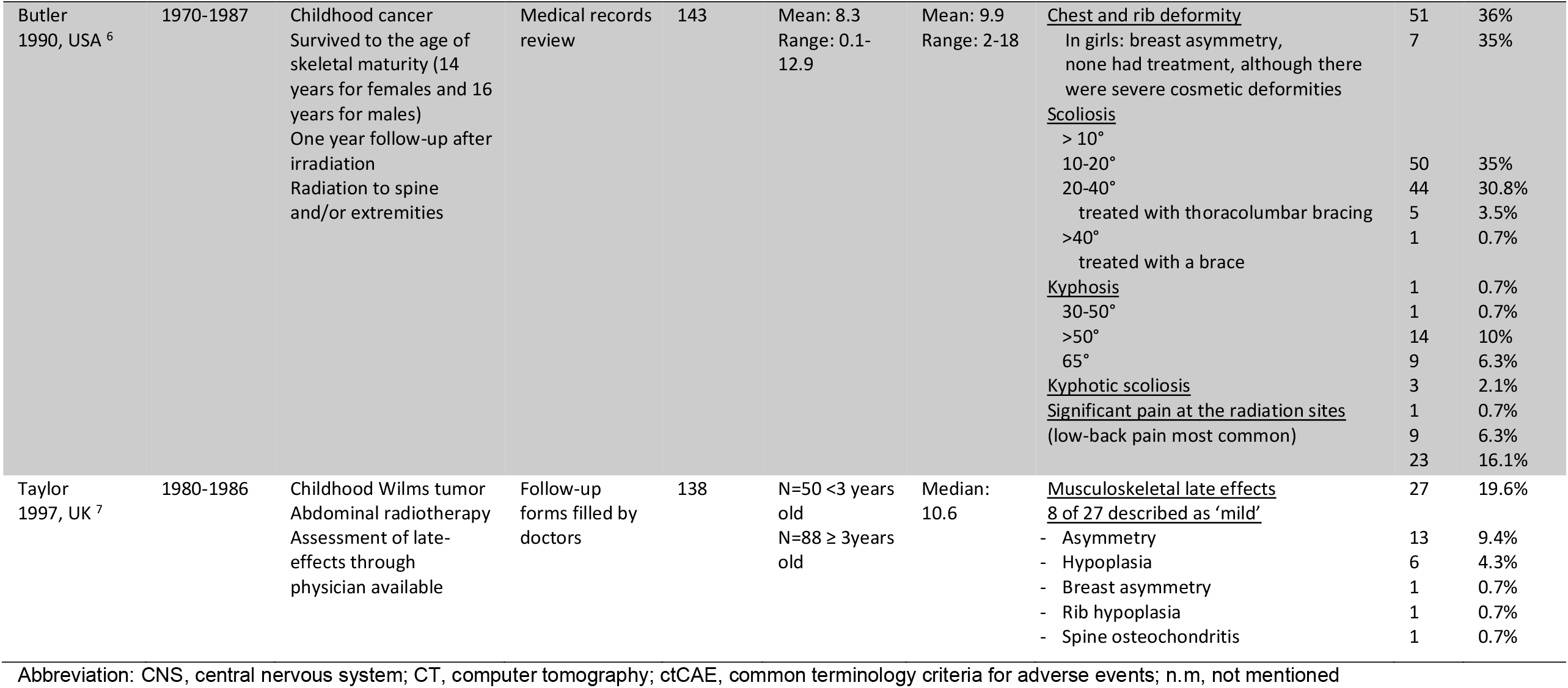
Literature summary of systematic review on chest wall abnormalities in childhood cancer survivors

Scoliosis was reported mainly in survivors of neuroblastoma and Wilms tumor, and after radiotherapy of the spine^2-4,8,9^. Breast asymmetries with severe cosmetic deformities were described in 35% of female survivors after radiation to the spine^6^. Breast underdevelopment was reported in 11% of survivors who had undergone chest radiation.^5^ Scars resulting in psychological problems was described in 80% of survivors of solid malignancies after surgery.^5^ Asymmetries of the ribs, scapula and thorax were described after neuroblastoma, Wilms tumor, and chest and abdominal radiotherapy.^2,5,7^ Studies that focused on selected outcomes or treatments reported a higher prevalence of chest wall abnormalities than studies based on unselected survivor cohorts (table 4).

## 4. Discussion

This is the first study to describe self-reported chest wall abnormalities in an unselected, representative sample of childhood cancer survivors in detail, and to validate answers in a structured interview. Two percent of all survivors reported a chest wall abnormality. We found a broad range of problems that was summarized as chest wall abnormalities. More than half of interviewed survivors were affected in their daily lives and three quarters had required medical attention.

A strength of this study is, that we clarified types of chest wall abnormalities and their impact on daily life by directly interviewing survivors. Survivors could explain their problems and we could inquire about the impact of chest wall abnormalities. We were able to reach 80% of eligible survivors who had reported a chest wall abnormality in the SCCSS questionnaire. Participants did not differ from nonparticipants (results not shown) and a previous study concluded that response bias in the SCCSS did not markedly influence prevalence estimates.^18^

The prevalence of chest wall abnormalities in this study and in a previously published study from Switzerland with an overlapping population^12^ was slightly higher than in to the North American Childhood Cancer Survivors Study (2.0% versus 1.3%), especially for CNS tumor survivors (4.0% versus 0.4%).^11,12^ Treatment-related factors like the frequency and cumulative dose of thoracic radiotherapy, such as spinal radiation in CNS tumor patients, might differ between countries and help to explain such differences in prevalence of chest wall abnormalities. Smaller studies and case series reported chest wall abnormalities in selected subgroups of survivors and found a wide range of prevalences (table 3). ^1-8^

Risk factors for chest wall abnormalities in survivors vary between studies. Our study is the first to report older age at diagnosis (16-20 years) as a risk factor for chest wall abnormalities. Peak bone growth velocity and increase of peak bone mass happen during puberty,^19^ therefore cancer treatment during this vulnerable time may affect the development of the spine and thoracic wall more severely than treatment earlier in childhood. CNS tumor survivors were most likely to report chest wall abnormalities. CNS tumors are often treated with radiotherapy to the spine, which another risk factor identified in our study (OR 2.0; 95%CI 1.0 – 4.2), in the North American Childhood Cancer Survivor Study (rate ratio 5.0),^11^ and in other studies.^5-8^ Also, CNS tumor survivors often suffer from co-morbidities that include small stature, functional deficits, endocrine diseases, fatigue, and psychological problems,^20^ which might lead to a higher subjective burden by chest wall abnormalities compared to other survivor groups.

We could validate chest wall abnormalities in eighteen of 20 survivors who indicated a chest wall abnormality in the SCCSS questionnaire. Two survivors had scars only, which they reported as chest wall abnormalities and 12 had both, scars and chest wall abnormalities. This suggests that not all survivors understood the term “chest wall abnormality” as was intended by the questionnaire. We suggest that future questionnaires describe chest wall abnormalities in more detail or use open questions to further assess the type of chest wall abnormality. For physicians involved in follow-up care of childhood cancer survivors awareness of chest wall abnormalities should be raised and clinical examinations of performed to quantify the extent of individual problems and limitations.

Many participants were affected in daily life by chest wall abnormalities, which reflects the severity of this rare late effect after childhood cancer treatment. The most common complaints impaired flexibility and physical fitness. An interdisciplinary treatment approach could help improve these issues. Early physiotherapy could be used in survivors at risk of developing chest wall abnormalities to improve late functional outcomes and might reduce pain.

## Conclusion

In conclusion, this study suggests that even though chest wall abnormalities are rare in the entire survivor population, they have a considerable impact on survivors lives. Physicians should pay close attention to these problems during follow-up care.

## Data Availability

The datasets generated and/or analysed during the current study are not publicly available due to the local data safety agreement but are available from the corresponding author on reasonable request.

## Abbreviations

CI: Confidence interval
CNS: Central nervous system
IQR: Interquartile range
N: Number
OR: Odds ratio
P: P-value
SCCR: Swiss Childhood Cancer Registry
SCCSS: Swiss Childhood Cancer Survivor Study

## Acknowledgements

We thank all childhood cancer survivors and families for participating in our survey. We thank the study team of the SCCSS (Fabiën Belle, Rahel Kuonen, Jana Remlinger, Cornelia Rebholz, Corina Rueegg, Grit Sommer, Annette Weiss, Laura Wengenroth). We would like to thank Christopher Ritter for his editorial assistance.

