## Supplementary Material for "Chest wall abnormalities in Swiss childhood cancer survivors"

**Journal: MedRXiv**

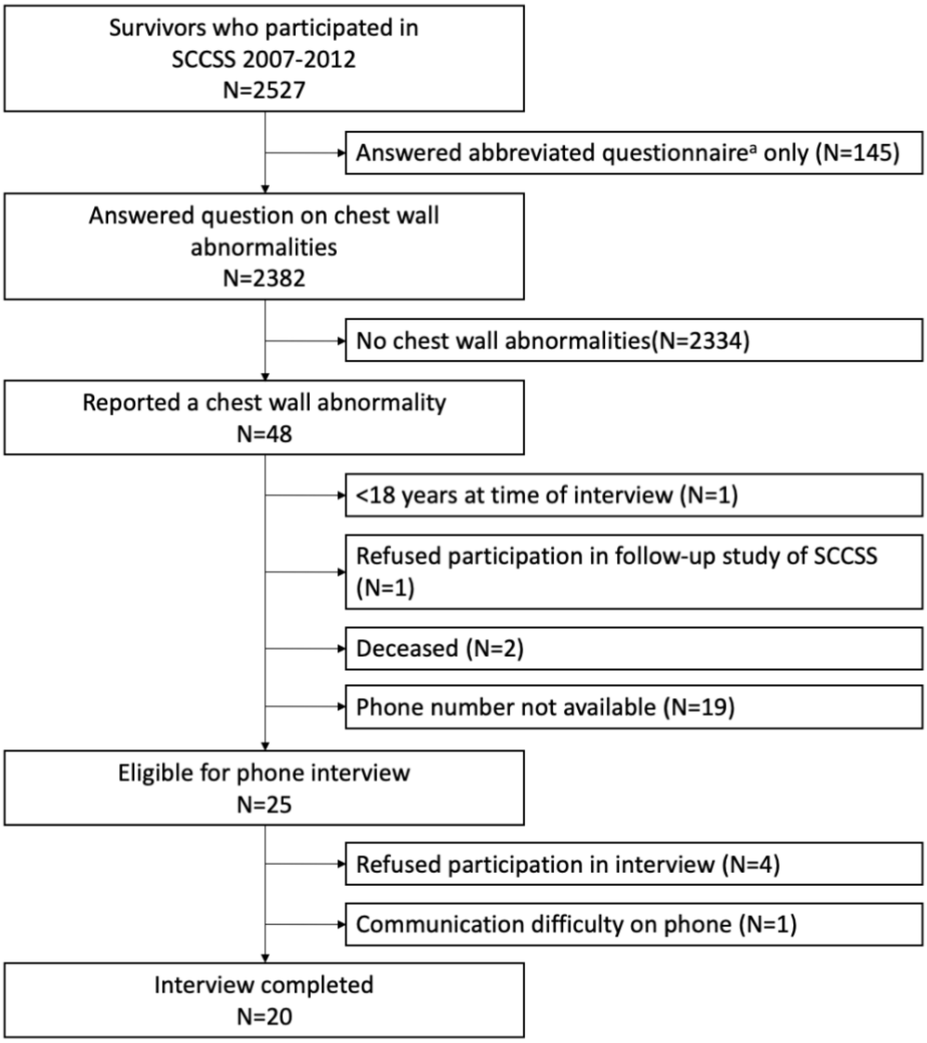


**Supplementary figure S1.** Study flowchart for the participation in the telephone interview on chest wall abnormalities in the Swiss Childhood Cancer Survivor Study.

[a] Addresses who did not complete the long questionnaire after mailings were offered an abbreviated version, which did not include the question on chest wall abnormalities
Abbreviations: SCCSS, Swiss Childhood Cancer Survivors Study; N, number

**A) Original question in German:**

| **Atmungssystem (Lungen)** | | | | | |
|---|---|---|---|---|---|
| | **Irgendwann im Leben** | | **Seit wann?** | **Aktuell noch vorhanden?** | |
| **Bitte kreuzen Sie an, ob ein Arzt Ihnen je mitgeteilt hat, dass sie folgendes haben…** | **Ja** | **Nein** | | **Ja** | **Nein** |
| Veränderungen am Brustkasten und/oder an den Rippen | ☐ | ☐ | ____ (Jahr) | ☐ | ☐ |

**B) Original question in French:**

| **Système respiratoire (Poumon)** | | | | | |
|---|---|---|---|---|---|
| | **Déjà eu?** | | **Quand?** | **Encore present?** | |
| **Veuillez cocher, si un médecin vous a déjà dit que vous aviez l'un des problèmes suivants…** | **Oui** | **Non** | | **Oui** | **Non** |
| Cage thoracique et/ou côtes anomale(s) | ☐ | ☐ | ____ (Année) | ☐ | ☐ |

**C) English Translation:**

| **Respiratory System (Lungs)** | | | | | |
|---|---|---|---|---|---|
| | **Ever in life?** | | **Since when?** | **Currently?** | |
| **Have you ever been told by a doctor that you have, or have had…** | **Yes** | **No** | | **Yes** | **No** |
| Changes on your thorax and/or ribs | ☐ | ☐ | ____ (Year) | ☐ | ☐ |

**Supplementary figure S2** Original question in A) German, B) French) and C) English translation of original question for adults on pulmonary health in the SCCSS questionnaire

### Supplementary text

#### Systematic literature review

#### Clinical questions:

What chest wall abnormalities can occur in childhood cancer survivors as a late effect of childhood cancer and childhood cancer treatment?

What is the prevalence of chest wall abnormalities in childhood cancer survivors?

What are risk factors for chest wall abnormalities in childhood cancer survivors?

#### PICO question:

| | |
|---|---|
| **P (Population)** | childhood, adolescent, and young adult cancer survivors |
| **I (Intervention)** | n/a |
| **C (Comparison)** | n/a |
| **O (Outcome)** | chest wall abnormalities |

#### Search terms:

| **Search 1: Population Cancer** | |
|---|---|
| **Textwords:** | (Cancer* [Title/Abstract] OR Tumor [Title/Abstract] OR Tumors [Title/Abstract] OR Tumour [Title/Abstract] OR Tumours [Title/Abstract] OR carcinom*[Title/Abstract] OR neoplas*[Title/Abstract] OR malignan*[Title/Abstract] OR leukemia [Title/Abstract] OR leukaemia [Title/Abstract] OR lymphom*[Title/Abstract] OR hematopoietic stem cell transplantation [Title/Abstract] OR bone marrow transplant*[Title/Abstract])<br><br>**OR** |
| **MESH:** | "Neoplasms"[Mesh] OR "Hematopoietic Stem Cell Transplantation"[Mesh] OR "Bone Marrow"[Mesh] |
| **Search 2: Population Children/Adolescents** | |
| **Textwords:** | (child* [Title/Abstract] OR infan*[Title/Abstract] OR adolescen*[Title/Abstract] OR pediatric*[Title/Abstract] OR paediatric*[Title/Abstract] OR young adult*[Title/Abstract] OR juvenil*[Title/Abstract])<br><br>**OR** |
| **MESH:** | ("Child"[Mesh] OR "Infant"[Mesh] OR "Adolescent"[Mesh] OR "Pediatrics"[Mesh] OR "Young Adult"[Mesh]) |
| **Search 3: Population Survivors** | |
| **Textwords:** | (Surviv*[Title/Abstract] OR follow-up[Title/Abstract] OR followup[Title/Abstract] OR long-term[Title/Abstract] OR following treatment* [Title/Abstract] OR after treatment* [Title/Abstract] OR post treatment*[Title/Abstract] OR treatment complet*[Title/Abstract] OR treatment termina*[Title/Abstract] OR late effect*[Title/Abstract] OR treatment related[Title/Abstract] OR sequela*[Title/Abstract] OR treatment induced[Title/Abstract] OR after diagnosis[Title/Abstract] OR retrospective[Title/Abstract] OR correlation [Title/Abstract]) |

| | |
|---|---|
| **MESH:** | **OR**<br><br>("Survivors"[Mesh] OR "Cancer Survivors"[Mesh] OR "Survival"[Mesh] OR "Aftercare"[Mesh]) |
| **Search 4: Outcome Chest wall abnormalities** | |
| **Textwords:**<br><br><br><br><br>**MESH:** | (Chest wall abnorm* [Title/Abstract] OR abnormal chest wall [Title/Abstract] OR Chest wall deform* [Title/Abstract] OR Chest wall asymmetr* [Title/Abstract] OR (chest wall[Title/Abstract] AND growth disturbance*[Title/Abstract]) OR (chest wall[Title/Abstract] AND growth retardation*[Title/Abstract]) OR Chest wall resection* [Title/Abstract] OR Kyphosis [Title/Abstract] OR Kyphotic deformity [Title/Abstract] OR Scoliosis [Title/Abstract] OR Scoliotic deform* [Title/Abstract] OR Lordosis [Title/Abstract] OR Thoracic wall deform* [Title/Abstract] OR Thoracic wall abnorm* [Title/Abstract] OR (Thoracic wall[Title/Abstract] AND asymmetr*[Title/Abstract]) OR (Thoracic wall[Title/Abstract] AND growth disturbance*[Title/Abstract]) OR (Thoracic wall[Title/Abstract] AND growth retardation[Title/Abstract]) OR Rib asymmetry* [Title/Abstract] OR Rib deform* [Title/Abstract] OR Rib abnorm* [Title/Abstract] OR Spinal deform* [Title/Abstract] OR Spinal abnorm* [Title/Abstract] OR Spinal asymmetry* [Title/Abstract] OR (Spinal*[Title/Abstract] AND growth disturbance[Title/Abstract]) OR (Spinal*[Title/Abstract] AND growth retardation[Title/Abstract]) OR Spinal malalignment [Title/Abstract] OR Spinal hypoplasia [Title/Abstract] OR Spinal deform* [Title/Abstract] OR (Reduction*[Title/Abstract] AND Spinal growth[Title/Abstract]) OR Impaired vertebral growth [Title/Abstract] OR Vertebral asymmetr* [Title/Abstract] OR Vertebral abnorm* [Title/Abstract] OR Vertebral malformation* [Title/Abstract] OR (Alternation[Title/Abstract] AND axial alignment[Title/Abstract]) OR Pectus carinatum [Title/Abstract] OR Pectus excavatum [Title/Abstract] OR Sternal defect* [Title/Abstract] OR Sternal deform* [Title/Abstract] OR Sternal malformation* [Title/Abstract] OR sternal abnorm* [Title/Abstract] OR ((musculoskeletal[Title/Abstract]) AND (adverse effect*[Title/Abstract] OR abnorm*[Title/Abstract] OR late effect*[Title/Abstract] OR sequela*[Title/Abstract] OR asymmetr*[Title/Abstract] OR deform*[Title/Abstract] OR growth disturbance*[Title/Abstract] OR growth retardation*[Title/Abstract] OR hypoplasia*[Title/Abstract] OR disease*[Title/Abstract] OR impairment*[Title/Abstract])) OR (asymmetric [Title/Abstract] AND vertebral growth [Title/Abstract]))<br><br>OR<br><br>"Thoracic Wall"[Mesh] OR "Musculoskeletal System"[Mesh:NoExp] |
| **Search 1 AND 2 AND 3 AND 4**<br><br>**Publication date from 1975/01/01 to 2019/12/31;**<br>**English; French; German**<br>**Humans only** NOT (animals[mh] NOT humans[mh])<br>**Exclude publications types:** NOT (letter[pt] OR news[pt] OR comment[pt] OR editorial[pt] OR congresses[pt] OR abstracts[pt]) | |

### Search results:

| **Search platform** | **Before deduplication** | **After deduplication** |
|---|---|---|
| PubMed | 1228 | 1224 |
| Embase Ovid | 1238 | 695 |
| Google Scholar | 300 | 248 |
| **Total** | **2766** | **2167** |

599 duplicate records have been removed

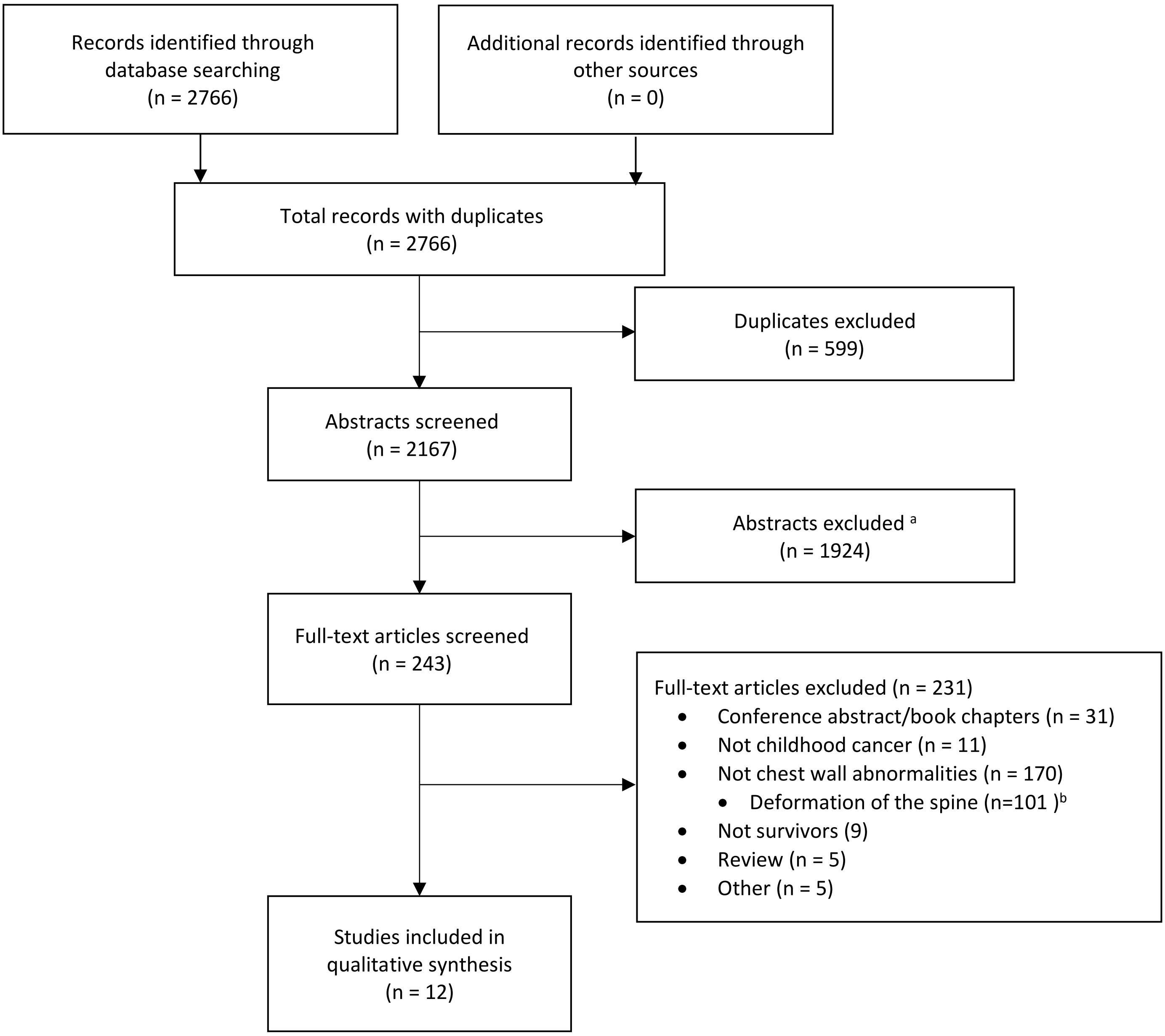


**Supplementary figure S3**. Flow diagram of article screening process

[a] Inclusion criteria for abstract screening: Study population: survivors of childhood, adolescent and young adult cancer (>=50% of population diagnosed prior to age 20 years); outcome: chest wall abnormality; all study designs; published 1975 onwards. Languages that were not spoken by members in the group (included are English, French, and German)
[b] Full texts focusing on deformation of the spine (kyphosis, kyphoscoliosis, scoliosis) only were excluded.
